# Activation status of immune cells in the airway is a defining feature of severe fungal asthma

**DOI:** 10.1101/2025.05.14.25327430

**Authors:** Emily L Plumpton, Stefano AP Colombo, Matthew Steward, Sheila L Brown, Saba Khan, Gaël Tavernier, Helen Francis, Hazel Platt, Tracy Hussell, William Horsnell, David Denning, Robert Niven, Angela Simpson, Andrew S MacDonald, Peter C Cook

## Abstract

Airborne fungi are potent inducers of respiratory disease and cause the debilitating conditions severe asthma with fungal sensitisation (SAFS) and allergic bronchopulmonary aspergillosis (ABPA). However, the immune cell types and the inflammatory airway environment that defines SAFS and ABPA patients is not extensively characterised. To address this, we recruited SAFS and ABPA patients, asthmatics without evidence of fungal sensitisation and healthy controls (n= 20 individuals per group). Immune cells were isolated from collected sputum and peripheral blood samples and immunophenotyping was performed via flow cytometry. By applying a machine learning approach to our dataset, we identify a critical association between CD4^+^ T cells, type 2 conventional dendritic cells, eosinophils, proinflammatory factors and severe respiratory disease. These complex immune signatures should be investigated further to improve the diagnostics and treatment of SAFS and ABPA.

**Graphical abstract:** 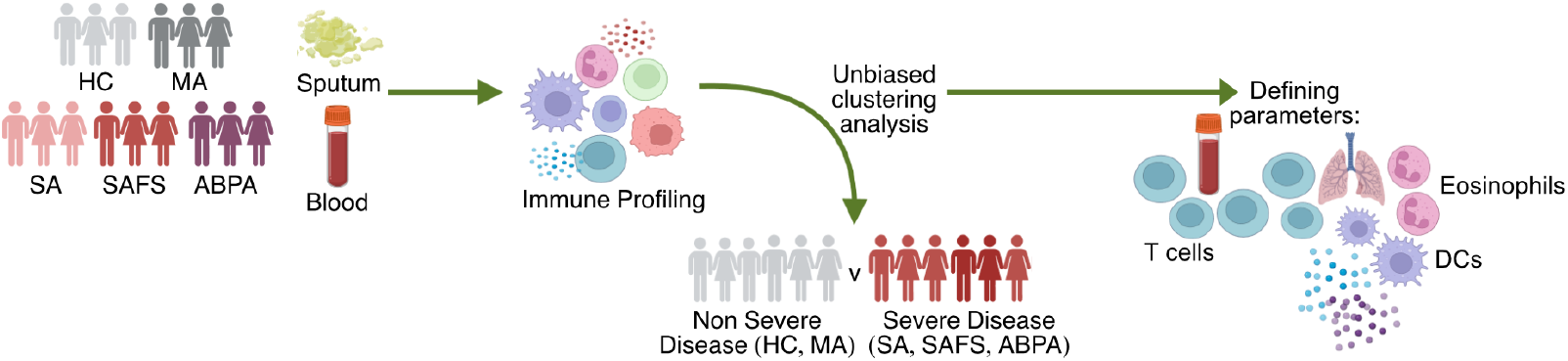

## Introduction

Global estimates of asthma prevalence are rising, with an estimated 262 million affected individuals in 2019 (1). Purported reasons for this rapid rise include urbanisation of lower- and middle-income countries resulting in higher rates of obesity, poorer air quality, diminished early exposure to pathogens, and improved asthma diagnostics (2–4). For most asthmatics, symptoms can be controlled with inhaled therapies. However, in 10% of patients (severe asthma SA patients), high doses of inhaled corticosteroid (ICS) and a second controller medication are required to prevent exacerbations. Even then, their asthma may remain poorly controlled (5, 6). Variation in treatment response can be attributed in part to the different types of inflammation which underpin asthma. Between 70-80% of patients have type 2 (T2) inflammation which is characterised by eosinophilia and the T2 cytokines IL-4, IL-5 and IL-13 (7–11) These patients are generally responsive to oral corticosteroids (OCS) and novel biologic therapies such as Benralizumab (anti-IL-5 receptor), Mepolizumab and Reslizumab (anti-IL-5), Omalizumab (anti-IgE), or Dupilumab (anti-IL-4/IL-13 receptor) (11). However, a significant proportion of patients have T2 ‘low’ asthma, the majority of whom experience airway neutrophilia and elevated levels of IL-8, IL-17 and IL-22. These patients are often less responsive to conventional treatments and less is known about this form of the disease (9, 10, 12).

Fungi such as *Aspergillus fumigatus (Af), Alternaria alternata* and *Cladosporium herbarum*, can be potent inducers and exacerbators of respiratory disease (13–17). In severe asthma, 30-50% of patients are sensitive to fungi, and this is strongly correlated with poor treatment response, reduced lung function, increased hospitalisation and asthma-related deaths (16, 18). Clinically, this is either described as severe asthma with fungal sensation (SAFS) or allergic bronchopulmonary aspergillosis (ABPA) (19, 20). Despite effecting 10 million people worldwide, SAFS and ABPA remain challenging to diagnose and treat (14, 21). The current diagnostic criteria for SAFS are 1) the presence of severe asthma, 2) elevated serum total IgE (<1000IU/mL) 3) elevated specific IgE (>0.35kU/L) to fungal allergens and/or a positive skin-prick to test to fungal allergens 4) the absence of clinical/radiological features of bronchiectasis. For ABPA diagnosis, total IgE must exceed 500 IU/mL and two additional criteria are required. Such include radiological evidence of bronchiectasis, blood eosinophilia >0.5 x 10^9^/L, or positive *Aspergillus* precipitins/IgG (19, 20). However, it is likely that a large proportion of ABPA patients, particularly in the developing world, are misdiagnosed and treated for recurrent bacterial infections (22, 23). Therefore, it is imperative to improve diagnostic testing to allow subsequent appropriate treatment strategies for SAFS and ABPA.

Much of our mechanistic understanding of fungal sensitisation in the lung comes from murine models (17, 24). Our group has recently shown that dendritic cell (DC) recognition of early growth stages of *Af* spores plays an important role in T cell mediated allergic airway inflammation, with the capacity to induce both eosinophilia and neutrophilia (17). However, whether DCs and T cells are important contributors in driving fungal asthma in humans is poorly understood.

To better understand the events that underpin SAFS and ABPA, we characterised immune cell populations of sputum and matched blood samples from a large asthma cohort. Despite the technical challenges in isolating immune cells from sputum, we successfully utilised multi-parameter flow cytometry to identify the proportion and activation status of granulocytes, myeloid and T cell populations. With the application of machine learning approaches on cellular data, we found that severe asthma groups (regardless of fungal sensitisation) clustered separately from mild asthma (MA) and healthy controls (HC). However, the cellular parameters which were most informative in clustering patients varied between sputum and blood. Clustering from blood samples was based largely on the abundance and activation of CD4^+^ T cells. In contrast, clustering from sputum samples was informed by CD4^+^ T cells, DC subsets (particularly cDC2s) and eosinophils. Furthermore, we found that the concentration of IFN*γ*, IL-9 and eotaxin in sputum supernatants were also reliable indicators of asthma severity. These results show the importance of charactering the airway environment of asthma patients to better understand the immune events which cause disease. They further highlight CD4^+^ T cells and cDC2s as critical players in severe respiratory disease and potential risk factors of fungal sensitisation.

## Results

The immune events that occur in the airways of asthma patients, particularly those who are fungal sensitised, are poorly understood (21). To redress this, we isolated cells from the sputum and blood of HC, MA, SA, SAFS and ABPA patients (Fig. 1A).

**Figure 1.**
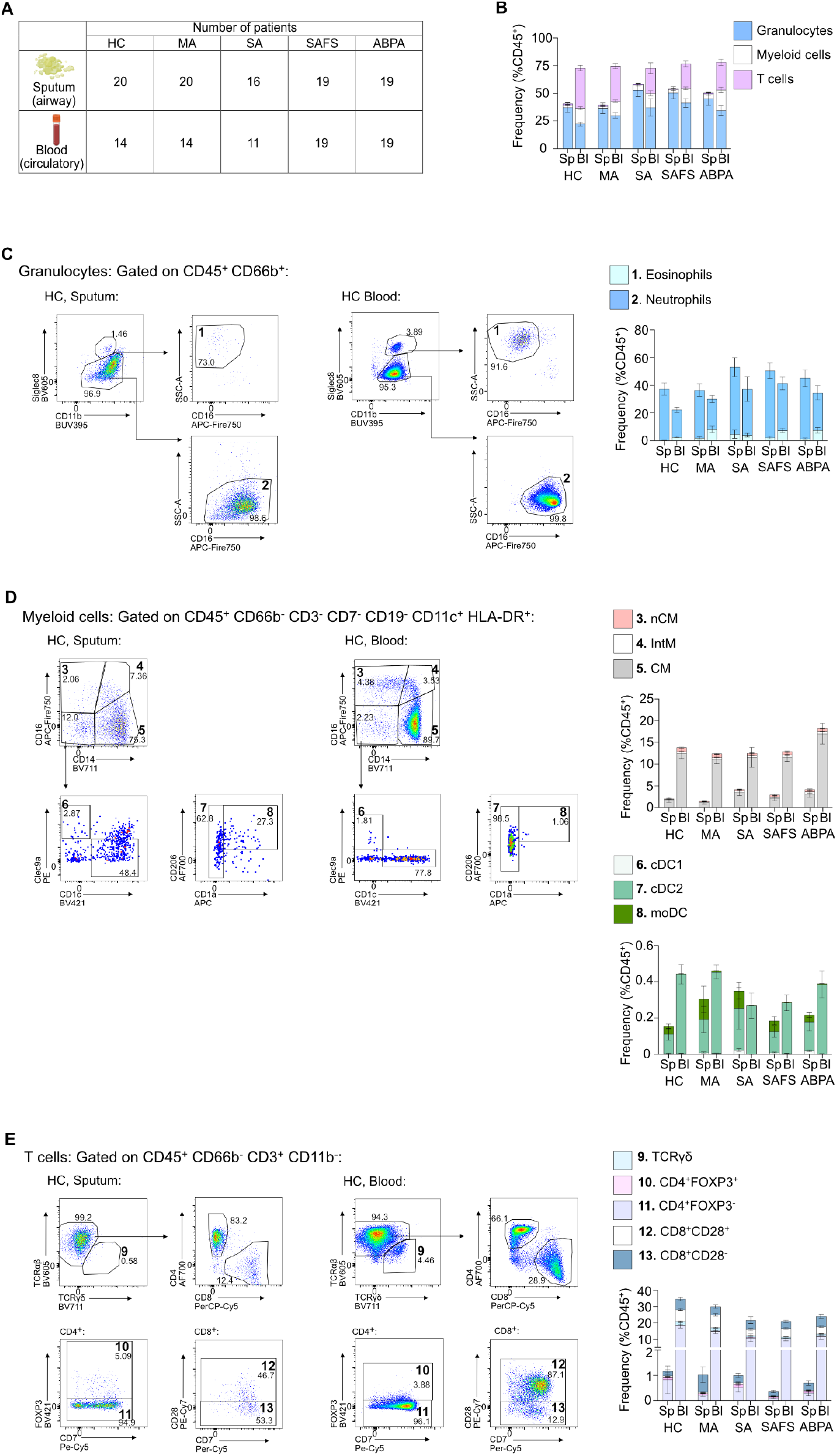
Identification of granulocyte, T cell and myeloid populations in the sputum and blood of severe asthma patients. **A)** Sputum and blood samples were collected from healthy controls (HC), mild asthmatics (MA), severe asthmatics (SA), severe asthmatics with fungal sensitisation (SAFS) and allergic bronchopulmonary aspergillosis (ABPA) patients. **B)** Frequency of granulocyte, T cell and myeloid populations in sputum (Sp) and blood (Bl), as percentage of CD45^+^ cells. Error bars show standard error of the mean. **C)** Flow plots to show the gating of eosinophils (1, CD66b^+^Siglec8^+^SSC^hi^CD16^lo^) and neutrophils (2, CD66b^+^Siglec8^-^ SSC^lo^CD16^hi^) in the sputum and blood of a HC. Stacked bar chart shows the frequency of eosinophils and neutrophils as % of CD45^+^ cells. Error bars show standard error of the mean. Representative flow plots to show the gating of non-classical monocytes (nCM) (3, CD11c^+^HLA-DR^+^CD14^-^CD16^hi^), intermediate monocytes (IntM) (4, CD11c^+^HLA-DR^+^CD14^hi^CD16^hi^), classical monocytes (CM) (5, CD11c^+^HLA-DR^+^CD14^hi^CD16^-^), type 1 conventional dendritic cells (cDC1) (6, CD11c^+^HLA-DR^+^CD14^-^CD16^-^CLEC9a^+^), type 2 conventional dendritic cells (cDC2) (7, CD11c^+^HLA-DR^+^CD14^-^CD16^-^CD1c^+^) and monocyte derived dendritic cells (moDC) (8, CD11c^+^HLA-DR^+^CD14^-^CD16^-^CD1c^+^CD1a^+^) in the sputum and blood of a HC. Stacked bar charts show the frequency of these monocyte and DC subsets as percentage of CD45^+^ cells. Error bars show standard error of the mean. **E)** Flow plots to show the gating of γ*δ* T cells (9, CD3^+^TCRγ*δ*^+^), CD4^+^ T cell populations (10, CD3^+^CD4^+^FOXP3^+^ and 11, CD3^+^CD4^+^FOXP3^+^) and CD8^+^ T cell populations (12, CD3^+^CD8^+^CD28^+^ and 13, CD3^+^CD8^+^CD28^-^) cells in the sputum and blood of a HC. Stacked bar chart shows the frequency of T cell subsets as percentage of CD45^+^ cells. Error bars show standard error of the mean.

The baseline characteristics of the study cohort are described in Table 1. Median age was 52 years (19-78years) and mean body mass index was 29.1±7.7 (kg/m^2^). Most participants were White British (65%) and never-smokers (57%). Available lung function data from our severe disease groups demonstrated significant airway obstruction. The mean forced expiratory volume in 1 second (FEV1)/forced vital capacity (FVC) ratio in SA, SAFS and ABPA patients was 0.65 ±0.12 to 0.63 ±0.14 and 0.58 ±0.13, compared to 0.84 ±0.03 in MA.

**Table 1:**
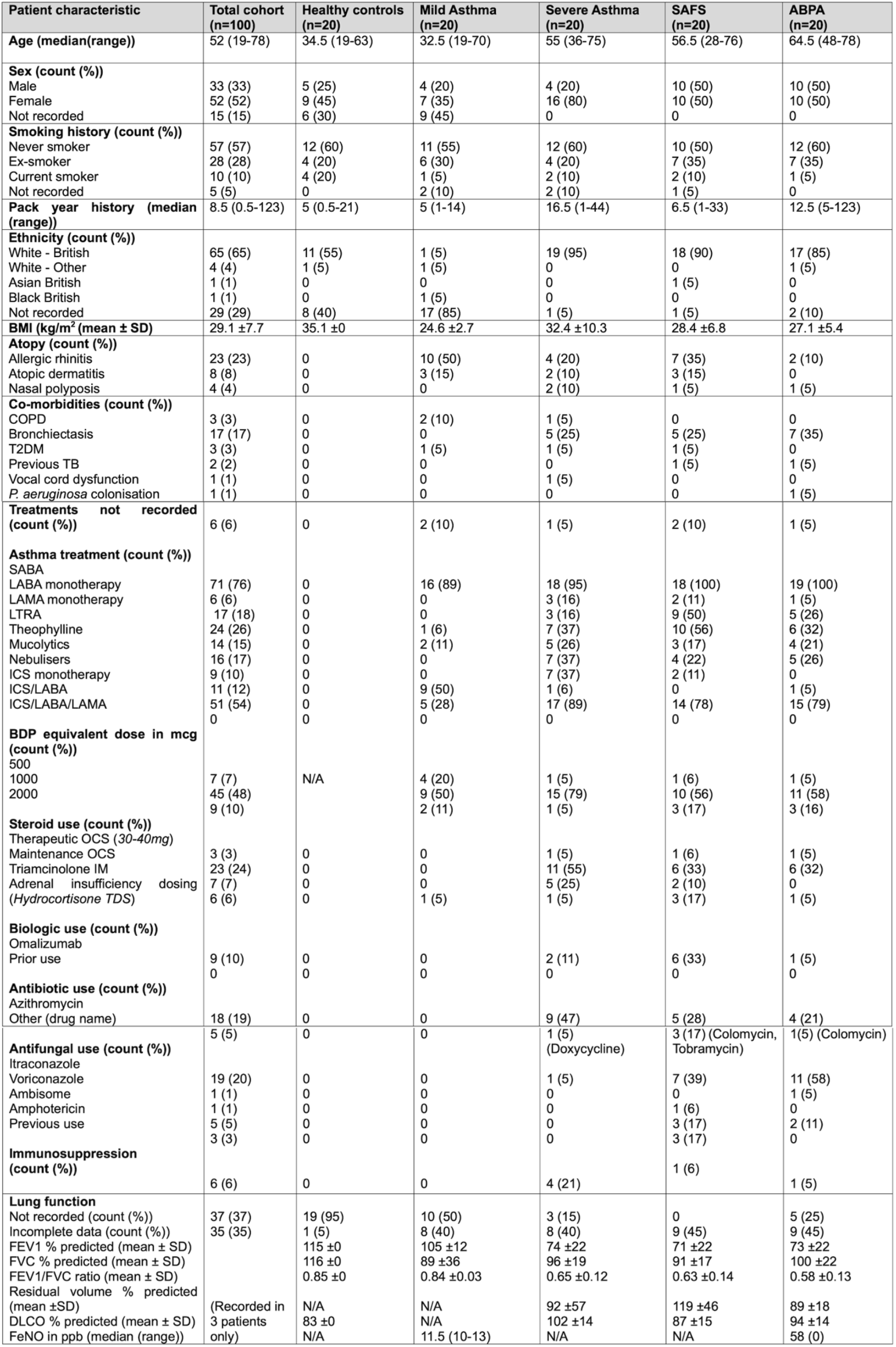
Patient cohort demographics and clinical characteristics. Abbreviations: SAFS, severe asthma with fungal sensitisation; ABPA, allergic bronchopulmonary aspergillosis; BMI, body mass index; SD, standard deviation; COPD, chronic obstructive pulmonary disease; T2DM, Tdiabetes mellitus; TB, tuberculosis; *P. aeruginosa, Pseudomonas aeruginosa*; SABA, short-acting β 2-agonist; LABA, long-acting β 2-agonist; LAMA, long-acting muscarinic antagonists; LTRA, leukotriene receptor antagonist; ICS, inhaled corticosteroid; BDP, Beclometasone dipropionate equivalent; OCS, oral corticosteroid; IM, intramuscular; TDS, thrice daily; ppb; parts per billion.

With recruitment taking place in 2018, this cohort would not have received recently approved biologic interventions such as Benralizumab, Mepolizumab and Dupilumab to control their asthma. Most patients were being treated with high dose ICS and additional controller medications. However, within the SA, SAFS and ABPA groups 11%, 33% and 5% of patients were taking the anti-IgE medication Omalizumab. Furthermore, whilst 47% of the SA cohort had been previously prescribed with the antibiotic Azithromycin, there was greater use of anti-fungals in the SAFS and ABPA patients (Table 1).

### The frequency of granulocytes, myeloid and T cell populations differ between sputum and blood, regardless of disease

Using a 26-parameter flow cytometry panel, we identified granulocytes, myeloid, and T cell populations in both sputum and blood samples (Fig. 1B, Supplementary Fig. 1, Supplementary Table 1). Following removal of highly autofluorescent cells (likely macrophages (25)), revealed the majority of CD45^+^ cells in the sputum were granulocytes (Fig. 1B). Though neutrophils dominated this compartment (Fig. 1C), eosinophils were also detected (Fig. 1C). In the blood, there was a more equal representation of granulocytes, T cells and myeloid cells (Fig. 1B). The frequency of eosinophils was largely consistent in blood and sputum across patient groups (Fig. 1C).

Within the myeloid and lymphoid families, we then identified classical, intermediate and non-classical monocytes, conventional DC and monocyte-derived DC subsets (Fig. 1D), as well as T-cell receptor (TCR)γδ, CD4^+^, CD8^+^ and regulatory T cell populations (Fig. 1E). Regardless of patient group, the proportion of these populations differed greatly between the sputum and blood (Fig. 1D & E). For example, TCR*γδ* cells were abundant in the blood, but largely absent from the sputum (Fig. 1E).

Having determined the frequency of these immune cell populations in both sputum and blood, the expression of various surface markers in key populations was assessed to elucidate function and activation state (Supplementary Table 2).

### Phenotyping immune cells in the blood identifies individuals with severe asthma

To determine whether the cytometric data could unbiasedly predict patient condition, we employed random forests (RF) machine learning, a supervised classification method which generates decision trees to cluster variables. We applied RF to blood and sputum data separately, to highlight whether one sample type was more accurate in identifying patients with severe asthma and fungal airway disease.

First, we utilised 34 parameters relating to the blood which described immune cell population frequency (measured as both percentages of total CD45^+^ cells and the direct parent population) and expression of markers associated with activation in certain immune cell populations (Supplementary Table 3). Clinical co-variates and data obtained from the sputum were not included. Our RF analysis revealed two clusters: HC and MA patients (defined as ‘non-severe’), and SA, SAFS and ABPA patients (defined as ‘severe’) (Fig. 2A & B). Despite heterogeneity across the cohort, the grouping of patients into ‘non-severe’ and ‘severe’ disease groups was significant, with an estimated error rate of 20% (Fig. 2B & C).

**Figure 2.**
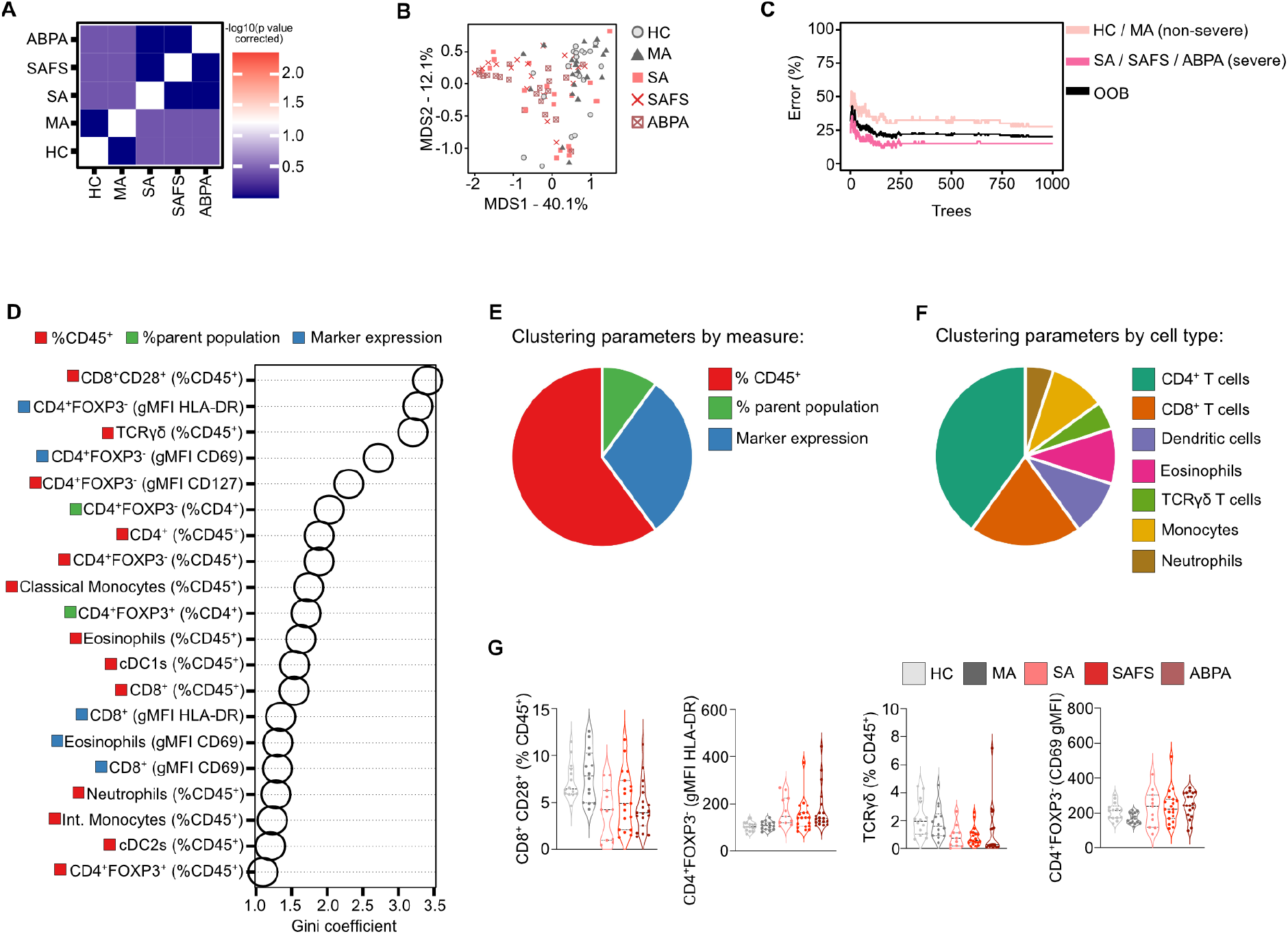
T cell abundance and activation in peripheral blood identifies severe asthma patients. Multivariate statistical analysis was performed on cellular immune parameters quantified by flow cytometry on cells isolated from peripheral blood. **A)** Tile plot representing the results of pairwise multivariate statistical comparisons between groups via Permutational MANOVA. **B-F)** Random forests (RF) analysis revealed: **B)** Classical multi-directional scaling ordination plot calculated from sample proximity within the RF model. **C)** Line graph representing the out-of-bag error rate of the RF model with increasing number of trees. **D)** The top 20 most informative parameters for guiding patient clustering, ranked via Gini coefficient. Red squares indicate parameters relating to population frequency as a percentage of CD45^+^ cells, green squares indicate parameters relating to population frequency as percentage of parent population, and blue squares indicate parameters relating to expression of markers that indicate cellular expression. **E)** Pie chart show the 20 most informative parameters, characterised by measure. **F)** Pie chart show the 20 most informative clustering parameters, categorised by cell type. **G)** Violin plots show the top four most informative clustering parameters. Each dot represents an individual patient and quartiles are shown by horizontal lines.

### Peripheral T cells discriminate between healthy individuals, mild disease inidviduals and individuals with severe airway disease

To highlight the cellular parameters which were most important for grouping patients, we extracted Gini coefficients from our RF model. Of the top 20 most significant clustering parameters (Fig. 2D), 15 described the frequency of an immune cell population (either as a percentage of CD45^+^ cells or the parent population) (Fig. 2E). This suggests that cumulatively, the abundance of immune cell populations in the blood was informative in deciphering asthma severity.

We then categorised the informative clustering parameters based on cell type and found that >60% related to T cells (CD4^+^, CD8^+^ and γ*δ* T cells) (Fig. 2F). Elevated abundance of CD4^+^FOXP3^-^ T cells and their increased expression of CD69, CD127, and HLA-DR (indicative of activated/antigen experienced T cells(refs (26, 27) (28, 29))), in the blood of severe groups (SA, SAFS, ABPA) indicates CD4^+^ T cells are more activated in these patients compared to non-severe groups (Fig. 2D-G). This agrees with preclinical work which found CD4^+^ T cells are crucial for allergic asthma (24, 30, 31). Furthermore, in alignment with previous studies (32, 33), the frequency of TCR*γ*δ cells and CD8^+^CD28^+^ T cells (CD28 being a co-stimulatory marker which is indicative of activation, maturity and antigen-experience (34)) were reduced in the blood of severe patients compared to the non-severe group (Fig. 2G).

Overall, these data highlight that the frequency and activation of peripheral T cell populations are major identifiers of patients with severe airway disease, regardless of fungal sensitisation.

### Sputum T cells, DCs and eosinophils provide robust identification of severe asthma patients

As performed on cells isolated from the blood, RF analysis of flow cytometry data on cells isolated from the sputum revealed two clear clusters of non-severe disease (HC and MA) versus severe disease (SA, SAFS and ABPA) (Fig. 3A & B). However, the difference between these two disease groups was greater when immunophenotyping cells isolated from the sputum compared to the blood (Fig. 2B-C & 3B-C).

**Figure 3.**
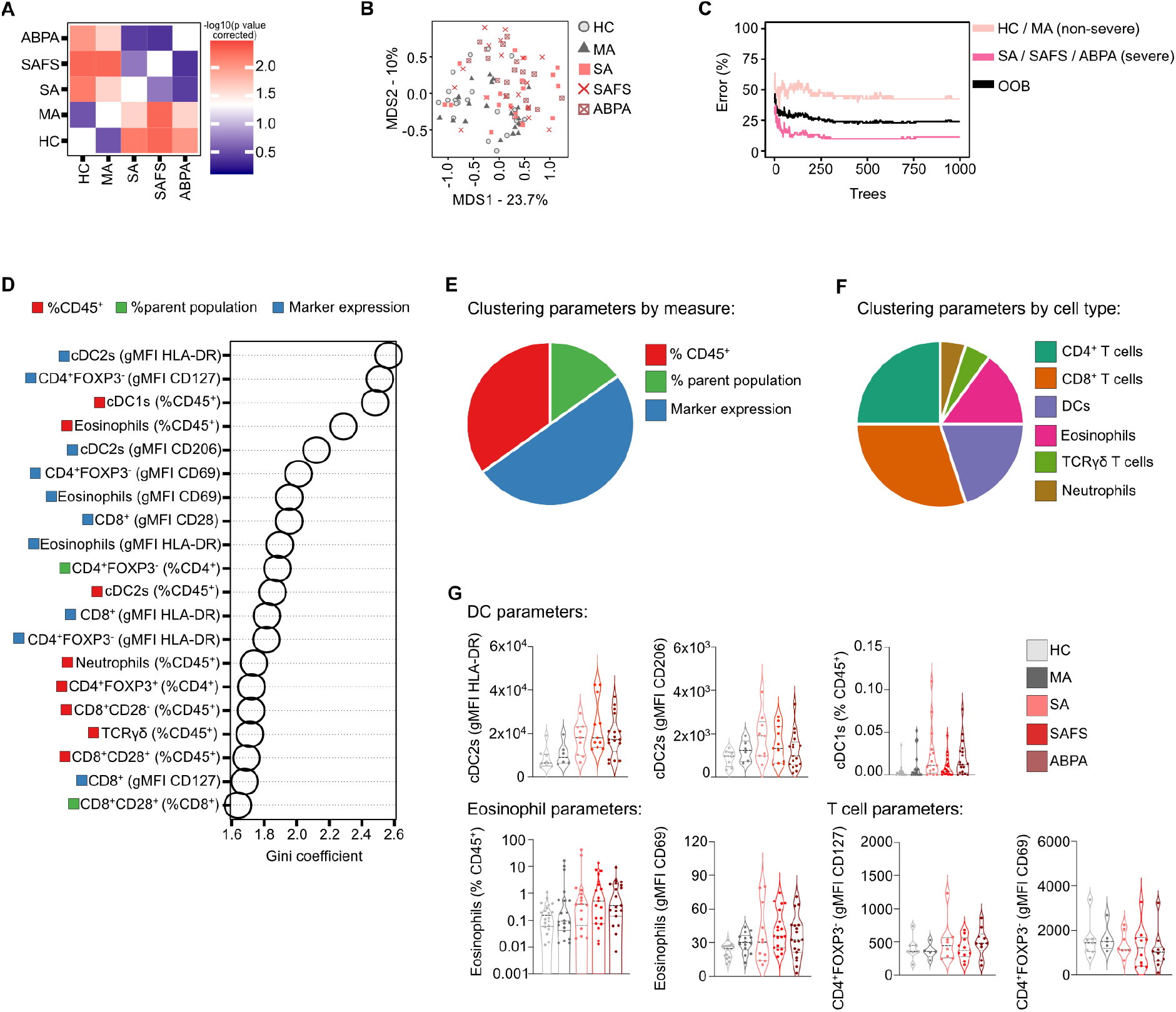
Abundance and activation of sputum T cells, dendritic cells and eosinophils identifies severe asthma patients. **A-F)** Multivariate statistical analysis was performed on cellular immune parameters quantified by flow cytometry on cells isolated from sputum. **A)** Tile plot representing the results of pairwise multivariate statistical comparisons between groups via Permutational MANOVA. **B-F)** Random forests (RF) analysis revealed: **B)** Classical multi-directional scaling ordination plot calculated from sample proximity within the RF model. **(C)** Line graph representing the out-of-bag error rate of the RF model with increasing number of trees. **(D)** The top 20 most informative parameters for guiding patient clustering, ranked via Gini coefficient. Red squares indicate parameters relating to population frequency as a percentage of CD45^+^ cells, green squares indicate parameters relating to population frequency as a percentage of parent population, and blue squares indicate parameters relating to cellular activation/ marker expression. **(E)** Pie chart to show the 20 most informative parameters, characterised by measure. **(F)** Pie chart to show the 20 most informative clustering parameters, categorised by cell type. **(G)** Violin plots to show the seven most informative clustering parameters. Each dot represents an individual patient and quartiles are shown by horizontal lines.

As seen in the blood, differences in the proportion of sputum CD4^+^ and CD8^+^ T cells were significant in guiding our model (Fig 3D-G). However, DC- and eosinophil-related parameters were also contributory (Fig. 3F) and activation state of immune cells was equally important to frequency in guiding clustering analysis (Fig. 3E). This supports characterisation of both abundance and activation of airway immune cell populations as important for appropriate understanding of disease immune-pathogenesis.

Despite DCs playing a central role in driving allergy, few studies have characterised DC populations in the sputum of asthma patients (35, 36). Here, we found cDC2 HLA-DR expression was the most informative airway readout (Fig. 3D) and was notably elevated on cDC2s isolated from patients with severe disease compared to HC (Fig. 3G). This indicates that DC activation and antigen presentation to T cells is increased in severe disease and could contribute to downstream inflammation (37–39). Similarly, the expression of the C-type lectin receptor CD206, previously associated with mature cDC2 populations in the lung (40), was elevated in the cDC2s of severe groups (Fig. 3G). cDC1s, another DC subset which may have a regulatory and homeostatic role in asthma (41), were also informative and elevated in the sputum of patients with severe asthma (Fig. 3D, 3G).

The frequency of airway eosinophils and their expression of the activation and maturation markers CD69 and HLA-DR (42–44), was also significant in unbiased patient clustering (Fig. 3D, 3G). This was expected as eosinophilia is often correlated with SA (7, 8, 45) and is widely used in asthma diagnostics (46). However, there was large variation in the proportion of sputum eosinophils within each disease group and this may be reflective of differences in treatment, co-morbidities and atopy status (Table 1).

### Secreted factors in the sputum, particularly IFNγ, IL-9 and eotaxin, were indicative of disease severity

Given that the cellular profile of the airway provided a strong indication of patient condition, we proceeded to quantify the concentration of 33 cytokines and secretory factors in sputum samples. Here pro-inflammatory, T2 and T17 cytokines/chemokines were measured, along with alarmins and growth factors. The majority of cytokines were more elevated in individuals with severe disease (Fig. 4A). Similarly to sputum eosinophilia, there was also considerable variation of secretory factors within groups (even in severe asthma patients) which may be due to differences in treatment regimens, co-morbidities or exacerbation status (47–51).

**Figure 4.**
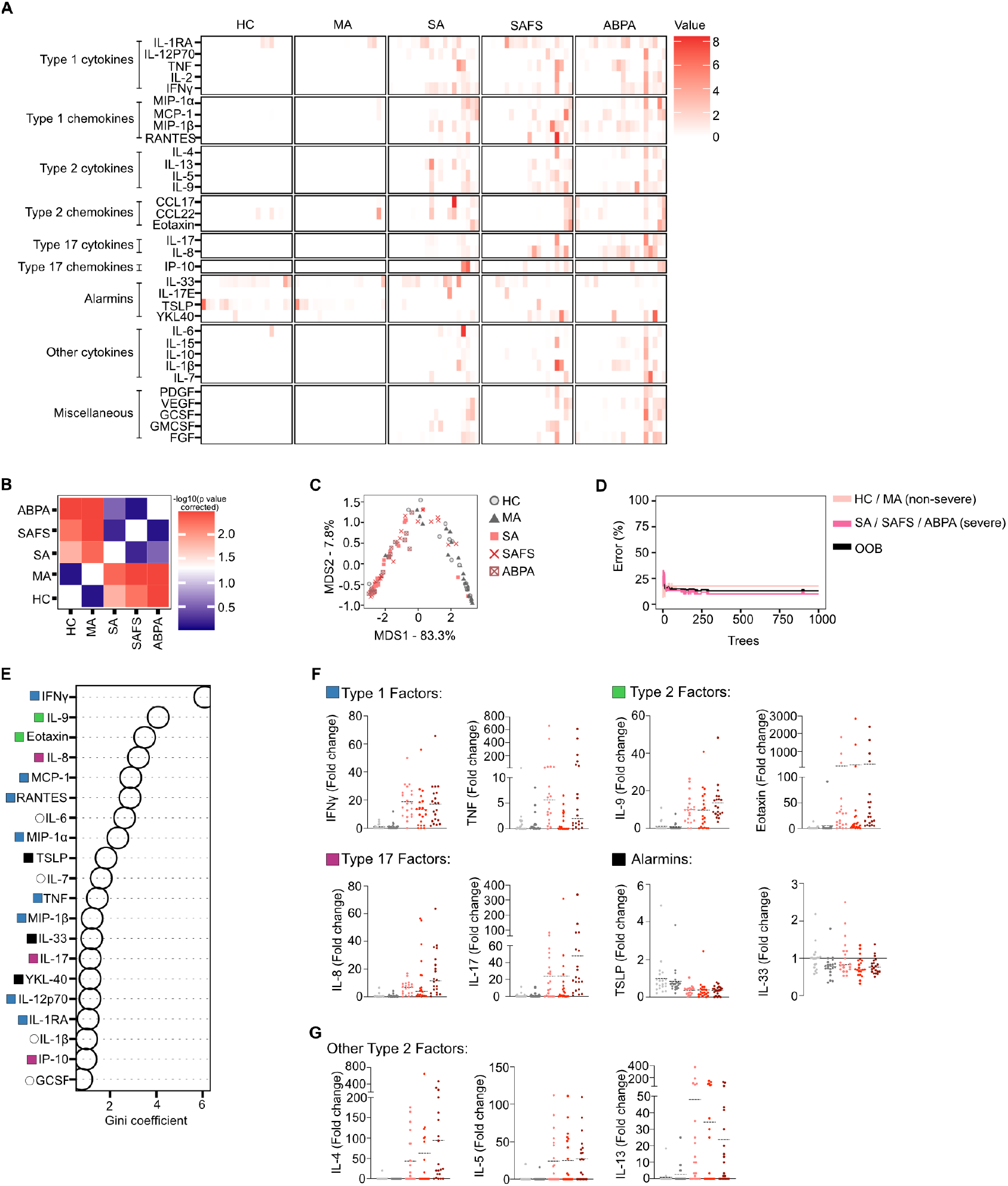
A range of secreted factors in the sputum predict severe asthma in a Random Forest analysis. **(A)** Heat map to show the concentration of secreted factors in each patient sputum sample, as measured by Luminex and ELISAs. **(B-E)** Multivariate statistical analysis was performed on these secreted factors. **(B)** Tile plot representing the results of pairwise multivariate statistical comparisons between groups via Permutational MANOVA. **(B-E)** Random forests (RF) analysis was performed using the randomForests package in R. **(C)** Classical multi-directional scaling ordination plot calculated from sample proximity within the RF model. **(D)** Line graph representing the out-of-bag error rate of the RF model with increasing number of trees. **(E)** The top 20 most informative secreted factors for guiding patient clustering, ranked via Gini coefficient. Blue squares indicate Type 1 factors, green squares indicate Type 2 factors, purple squares indicated Type 17 factors, black squares indicate alarmins and white circles indicate ‘other’. **(F)** Violin plots to show the most informative Type 1 factors (IFNγ, TNFα), Type 2 factors (IL-9, eotaxin), Type 17 factors (IL-8, IL-17) and alarmins (TSLP, IL-33), plotted as fold change, relative to HC mean. **(G)** Violin plots to show the concentrations of other Type 2 cytokines (IL-4, IL-5, IL-13). Each point represents an individual patient and dotted lines show the mean for each patient group.

Utilising the RF model on cytokine data, we found patients grouped into ‘non-severe’ or ‘severe’ disease with an estimated error rate of 13% (Fig. 4D). Thus, greater clustering accuracy was achieved than when the flow cytometry data was used. However, it was still hard to distinguish between HC and MA, as well as SA, SAFS and ABPA (Fig. 4B, 4C) using cytokine data alone.

IFN*γ* was the best predictor of disease, and was significantly increased in the airways of SA, SAFS and ABPA patients compared to HC (Fig. 4E, 4F). This pro-inflammatory cytokine is consistently found to be raised in the airway of asthmatics during viral exacerbation (49–51) and has also been associated with neutrophilic asthma (52). The T17 cytokines IL-8 and IL-17 were also ranked in the top 20 most influential secreted factors (Fig. 4E), both of which were elevated in the airways of patients with severe disease (Fig. 4F). Thymic stromal lymphopoietin (TSLP) and IL-33 (shown to be important drivers of asthma) were also deemed informative for clustering (Fig. 4E). However, in contrast to previous studies, TSLP and IL-33 were less concentrated in severe disease than non-severe disease (53, 54) (Fig. 4F). Notably, none of the T2 cytokines commonly used as biomarkers for eosinophilic asthma (IL-4, -5, -13), were ranked amongst the top 20 most informative clustering parameters (Fig. 4E). Nevertheless, they were still significantly elevated in severe disease groups compared to HC (Fig. 4G).

Taken together, these data support biomarkers signatures for severe fungal respiratory disease being complex and requiring further definition to confirm the potential for IFN*γ*, IL-9 and eotaxin as reliable indicators of disease type.

## Discussion

We collected paired blood and sputum samples from a large asthma cohort to define immune signatures that associate with SAFS and ABPA. Unbiased machine learning analysis of our datasets revealed the immune profile of HC and MA patients to be distinct from patients with SA, SAFS and ABPA. Furthermore, sputum samples, reflecting the airway, were more effective than blood samples for discriminating between these patient groups.

Due to difficulties associated with obtaining patient airway samples, most asthma research has been conducted on blood samples (55). Thus, many peripheral immune cell populations, including eosinophils, neutrophils, T cells, monocytes and DCs, have been shown to differ between healthly individuals and those with severe asthma (31). However, understanding which of these populations are most significant in driving disease is complex and likely to be ultimately driven by diverse contributory factors. Here, our RF analysis showed peripheral T cells to be the prominent population that distinguished between non-severe and severe airway disease, particularly CD4^+^ T cell activation and the frequency of TCR*γ*δ and CD8^+^CD28^+^ populations (Fig. 2D, F & G). This supports previous studies which have associated these factors with disease (32, 33, 56). Further, in the sputum, the expression of CD69 and HLA-DR on airway-associated CD4^+^FOXP3^-^ T cells was important in guiding clustering analysis (Fig. 1E). We suggest that these cells could be the source of the elevated IL-13 and IL-17 concentrations seen in the sputum of severe disease groups (Fig 4A & F). This would align with findings from our murine model of fungal allergic airway inflammation, which showed that IL-13^+^ and IL-17^+^ CD4^+^ T cells drive airway eosinophilia and neutrophilia (24). Our data identifies the importance of both systemic and local T cell activation in severe asthma, regardless of fungal sensitisation.

Further analysis of sputum samples revealed that in addition to T cells, DC subsets (cDC1s and cDC2s) could discriminate between MA/HC and SA, SAFS and ABPA individuals (Fig. 3D, 3E). Whilst the role of cDC1s in asthma is debated (41), previous studies in mouse models have shown that cDC2s are critical mediators of allergic inflammation to fungal and non-fungal allergens (24). In humans, a recent study on mild asthmatic patients revealed that airway cDC2s were key responders to allergen challenge (57) and here, we observed that their activation status helped to define patient condition (Fig 3D-G). To our knowledge, this study is the first to highlight the importance of cDC2 activation in the airways of SAFS and ABPA patients. However, future work is needed to focus on the functional capacity of cDC1s vs cDC2s from severe asthma patients and their contributions in causing disease.

In comparison to DCs, eosinophils are well characterised in human asthma and are used as a core diagnostic measure of T2 inflammation (46). In support of their clinical use, we found that sputum eosinophils were the fourth most important airway factor in identifying patients with severe disease (Fig. 3D). Their frequency also tended to be higher in SA compared to HC (Fig. 3G). Comparatively, blood eosinophils were less informative of disease severity as their frequency was ranked the 11^th^ most influential parameter for clustering analysis (Fig. 2D) However, this could be explained by the fact that all patients within our cohort were receiving treatment for their condition. 30% of patients were on corticosteroids (either therapeutically at 30-40mg, or as maintenance at ≤10mg) and 10% were receiving Omalizumab (Table 1), both of which are known to reduce peripheral eosinophil counts and control T2 inflammation (58–61). If samples had been collected prior to treatment, it is likely that eosinophilia would have been more pronounced in severe disease groups. The impact of recently-approved biologic therapies on the immune environment of these patients remains an important question to address.

As seen with the flow cytometry data, when RF analysis was applied to secreted factors from sputum supernatants, patients clustered into non-severe and severe disease groups (Fig. 4A-C). Out of the 34 secreted factors assessed, IFN*γ*, IL-9 and eotaxin were most important in predicting severe disease (Fig. 4E). These factors have been consistently associated with asthma, specifically IFN*γ* in the context of viral exacerbation (49–51) and eotaxin and IL-9 in the context of T2 inflammation and allergy (62–66). Overall, there is strong evidence to suggest that these cytokines should be considered for use in novel asthma diagnostics and treatments. Future work should aim to identify the cellular source of these cytokines and their importance in causing fungal sensitisation.

In summary, our work demonstrates how RF can identify key immune factors which distinguish health and disease. We found that cell populations and secreted factors in the sputum were much more effective indicators of severe airway disease than peripheral blood cells. However, regardless of sample type, there were no striking differences in the immune environment of SA, SAFS and ABPA patients. The mechanisms which underpin each specific condition could be revelated using more sensitive techniques such as single cell RNA-sequencing or *ex vivo* cell stimulation with fungal allergens. Further, the impact of novel biologic asthma therapies on such pathways remains to be defined. Nevertheless, our finding that sputum T cells, cDC2s and eosinophils as prominent features of SA could guide the development of improved diagnostics and treatments to improve SAFS and ABPA management.

## Material and methods

### Participant selection

Patients for this study were recruited from specialist clinics at the National Aspergillosis Centre in Manchester between January and June 2018. Paired blood and sputum samples were obtained alongside demographic data and clinical information. 20 patients were recruited to each of the following cohorts: healthy controls (HC), mild asthmatics (MA), severe asthmatics (SA), severe asthmatics with fungal sensitisation (SAFS), and allergic bronchopulmonary aspergillosis (ABPA) patients. HCs were defined as adults aged ≥18 with no significant past medical history or current prescription drug use. MAs were defined as those with objective evidence and a formal clinical diagnosis of asthma with persistent symptoms and use of reliever inhalers >2 days/week, but not daily. SAs were defined in keeping with the European Respiratory Society/American Thoracic Society definition of patients having uncontrolled disease despite use of high dose ICS and ≥1 additional controller medications (6). SAFS was defined as previously described and the presence of ABPA excluded (6). ABPA was defined as per International Society for Human & Animal Mycology (ISHAM) criteria at the time of recruitment (2018). All participants gave informed written consent. We acknowledge the Manchester Allergy, Respiratory and Thoracic Surgery Biobank for supporting this project and thank the study participants for their contribution, Rec: 20/NW/0302, IRAS: 285126 (North West - Haydock Research Ethics Committee).

### Sample collection and cell isolation

Sputum samples (spontaneous or induced by hypertonic saline) were collected and processed as previously described (67). Samples were centrifuged and supernatants and cells cryopreserved at −150ºC, with cells resuspended in freezing media (50% DMEM, 10% DMSO and 50% FBS (all Sigma) controlled freezing of sputum cells was obtained by use of a CoolCell Freezer container (Corning)).

Blood samples, taken at same time as sputum collection were collected in EDTA-coated tubes and transported to the laboratory at room temperature. Cells were resuspended in sterile water for 10 seconds, to lyse red blood cells and then cryopreserved in freezing media and stored at −150°C. Controlled freezing of cells isolated from sputum and blood was obtained by use of a Mr Frosty freezer container (Nalgene).

### Flow cytometry

Sputum and blood samples were thawed in a 37°C water bath and resuspended in pre-warmed RPMI + 10% FCS + DNase. Following centrifugation, the pellet was re-suspended in ice cold FACS (PBS containing 2% FBS and 2mM EDTA). Cells were washed with PBS and stained for viability with Live/Dead Blue™ (1:2000; ThermoFisher). Cells were then blocked with Human FcBlock (BioLegend) in FACS buffer, before being stained for surface markers at 4°C for 30min. Surface stain was then removed by washing the samples twice in FACS buffer. To fix, cells were then before resuspended in 1% paraformaldehyde (PFA) in PBS and incubated at room temperature for 10min. PFA was removed and cells were washed three times in eBioscience permeabilization buffer (ThermoFisher) for the staining of intracellular cytokines at 4°C for 60min. Antibody details can be found in Supplementary Table 1. Data was acquired on BD Fortessa and analysis was undertaken using FlowJo™ v10.0 Software (BD Life Sciences). Sentinel gating was used to maximise the number of populations that we could identify from our 16-colour panel. Briefly, markers which were not co-expressed by the same immune cell population were assigned the same fluorophore. For example, the monocyte marker CD16 and the B cell marker CD19 were both assigned APC/Fire750. Other markers which shared the same fluorophore were TCR*γ*δ and CD14, Siglec-8 and TCRab, FOXP3 and CD1c, CD4 and CD206, Clec9a and CD127, CD1a and CD28. Gating strategies and representative flow cytometry plots are shown in Supplementary Figure 1. Due to the high variation in sputum sample quality and cell count, immune cell populations were excluded from downstream analysis if event number was below the set threshold value (Supplementary Table 3.

### Luminex and ELISAs

To quantify the concentration of various soluble factors in sputum supernatants, a Bio-Plex Pro Human Cytokine 27-plex Assay (BioRad) was performed per manufacturers instructions. R&D DuoSet ELISAs kits were used to quantify the concentration of CCL17, CCL22, TSLP, IL-33, YKL40 and IL-17E according to the manufacturers instructions. Where concentration was below the limit of detection, a substitute value of 1×10^-7^ was used to enable subsequent fold change calculations.

### Statistical analysis

Statistical analysis was performed using Prism v9 (GraphPad) and R (version 4.4.1). Multivariate testing using Permutational MANOVA was performed using the ‘adonis.pair’ function from the ‘EcolUtils’ package (version 0.1) with p values corrected for the false discovery rate (fdr).

### Random forest analysis

Random forests analysis was performed in R (version 4.4.1) using the ‘randomForest’ package (version 4.7-1.1). Prior to analysis, missing data points were imputed using the ‘rfImpute’ function. For each model the number of trees was set to 1000 and mtry was set to the level which produced the lowest out of bag error. Dimensionality reduction was performed using classical multidimensional scaling (MDS) by generating a distance matrix from proximity values calculated by the Random forest model using the ‘dist’ function then running ‘cmdscale’.

## Data Availability

All data contributing to the conclusions of this manuscript are present in the manuscript and supplementary material. Raw data are available upon reasonable request to the corresponding authors.

## Author contributions

P.C.C. was responsible for conceptualisation. E.L.P., S.A.P.C, M.S., S.L.B., S.K. performed the experiments or undertook data analysis. H.F. T.H., G.T., A.S., A.S.M., were instrumental in the acquisition and processing of samples from the primary patient cohort. E.L.P., S.A.P.C., M.S., W.H., P.C.C. prepared the manuscript with input from all authors.

## Acknowledgements

This manuscript was supported by the North West Lung Centre Charity at Manchester University NHS Foundation Trust. The authors would like to acknowledge the Manchester Allergy, Respiratory and Thoracic Surgery (ManARTS) Biobank (REC: 20/NW/0302). In addition, we would like to thank the study participants for their participation. The views expressed are those of the author(s) and not necessarily those of the North West Lung Centre Charity, the NIHR, or the Department of Health and Social Care. PCC was supported by funding from University of Manchester Dean’s Prise Early Career Research Fellowship, Springboard Award (Academy of Medical Sciences, SBF002/1076) a Wellcome Trust Sir Henry Dale Fellowship (218550/Z/19/Z). PCC, ELP and MS were supported by the MRC Centre for Medical Mycology at the University of Exeter (MR/N006364/2 and MR/V033417/1), and the NIHR Exeter Biomedical Research Centre (NIHR203320). MS was supported by MRC Doctoral Training Grant MR/W502649/1. The views expressed are those of the author(s) and not necessarily those of the NIHR or the Department of Health and Social Care. MS was supported with an MRC Doctoral Training Grant (MR/W502649/1). The views expressed are those of the authors and not necessarily those of the NIHR or the Department of Health and Social Care. For the purposes of open access, the author has applied a Creative Commons Attribution (CC BY) licence to any Author Accepted Manuscript version arising.

## Conflicts of interest declaration

None

## Online Supplemental Figure Legends

**Supplementary Figure 1**. Gating strategy used to identify cell populations in both blood and sputum. All gates are shown in an example HC blood sample. Technical gates are shown for the matched sputum sample.

**Supplementary Figure 2**. Column charts to show the number of CD45^+^ cells, granulocytes, myeloid and T cells in sputum and blood, across patient groups. Each point represents an individual patient and error bars show standard error of the mean.

**Supplementary Table 1:** Table of flow cytometry antibodies and reagents.

**Supplementary Table 2:** Surface marker expression was quantified in certain cell populations by mean fluorescence intensity. Table to show the markers included in quantification.

**Supplementary Table 3:** Exclusion criteria and threshold values set for downstream gating of samples.

## Notes

### Competing Interest Statement

The authors have declared no competing interest.

### Author Declarations

The North West-Haydock Research Ethics Committee of the National Research Ethics Service Committee gave ethical approval for this work.

## References

1. Vos T, Lim SS, Abbafati C, et al. Global burden of 369 diseases and injuries in 204 countries and territories, 1990–2019: a systematic analysis for the Global Burden of Disease Study 2019. The Lancet 2020; 396(10258):1204–22. doi: 10.1016/S0140-6736(20)30925-9

2. Yasaratne D, Idrose NS, Dharmage SC. Asthma in developing countries in the Asia-Pacific Region (APR). Respirology 2023; 28(11):992–1004. doi: 10.1111/resp.14590

3. Wang Z, Li Y, Gao Y, et al. Global, regional, and national burden of asthma and its attributable risk factors from 1990 to 2019: a systematic analysis for the Global Burden of Disease Study 2019. Respiratory Research 2023;24(24). doi: 10.1186/s12931-023-02475-6

4. Varghese D, Ferris K, Lee B, et al. Outdoor air pollution and near-fatal/fatal asthma attacks in children: A systematic review. Pediatric Pulmonology. 2024; 59(5):1196–206. doi: 10.1002/ppul.26932

5. Wang E, Wechsler ME, Tran TN, et al. Characterization of Severe Asthma Worldwide. Chest. 2020; 157(4):790–804. doi: 10.1016/j.chest.2019.10.053

6. Chung KF, Wenzel SE, Brozek JL, et al. International ERS/ATS guidelines on definition, evaluation and treatment of severe asthma. European Respiratory Journal. 2014; 43(2):343–73. doi: 10.1183/09031936.00202013

7. Ricciardolo FLM, Sprio AE, Baroso A, et al. Characterization of T2-Low and T2-High Asthma Phenotypes in Real-Life. Biomedicines. 2021; 9(11):1684. doi: 10.3390/biomedicines9111684

8. Frøssing L, Silberbrandt A, Von Bülow A, et al. The Prevalence of Subtypes of Type 2 Inflammation in an Unselected Population of Patients with Severe Asthma. The Journal of Allergy and Clinical Immunology: In Practice. 2021; 9(3):1267–75. doi: 10.1016/j.jaip.2020.09.051

9. Hinks TSC, Levine SJ, Brusselle GG. Treatment options in type-2 low asthma. European Respiratory Journal. 2021; 57(1):2000528. doi: 10.1183/13993003.00528-2020

10. Jackson DJ, Aljamil N, Roxas C, et al. P48 The ‘T2-low’ asthma phenotype: could it just be T2-high asthma treated with corticosteroids? Thorax. 2018; 73(4). doi: 10.1136/thorax-2018-212555.206

11. Howell I, Howell A, Pavord ID. Type 2 inflammation and biological therapies in asthma: Targeted medicine taking flight. Journal of Experimental Medicine. 2023; 220(7). doi: 10.1084/jem.20221212

12. Peri F, Amaddeo A, Badina L, et al. T2-Low Asthma: A Discussed but Still Orphan Disease. Biomedicines. 2023; 11(4):1226. doi: 10.3390/biomedicines11041226

13. Denning DW, O’Driscoll BR, Hogaboam CM, et al. The link between fungi and severe asthma: a summary of the evidence. European Respiratory Journal. 2006; 27(3):615–26. doi: 10.1183/09031936.06.00074705

14. Denning D. Global incidence and mortality of severe fungal disease. The Lancet Infectious Diseases. 2024; 24(7). doi: 10.1016/S1473-3099(23)00692-8

15. Namvar S, Warn P, Farnell E, et al. Aspergillus fumigatus proteases, Asp f 5 and Asp f 13, are essential for airway inflammation and remodelling in a murine inhalation model. Clinical & Experimental Allergy. 2015; 45(5):982–93. doi: 10.1111/cea.12426

16. Zureik M. Sensitisation to airborne moulds and severity of asthma: cross sectional study from European Community respiratory health survey. BMJ. 2002; 325(7361):411-. doi: 10.1136/bmj.325.7361.411

17. Houlder EL, Gago S, Vere G, et al. Aspergillus-mediated allergic airway inflammation is triggered by dendritic cell recognition of a defined spore morphotype. Journal of Allergy and Clinical Immunology. 2025; 155(3):988–1001. doi: 10.1016/j.jaci.2024.10.040.

18. Black PN, Udy AA, Brodie SM. Sensitivity to fungal allergens is a risk factor for life-threatening asthma. Allergy. 2000; 55(5):501–4. doi: 10.1034/j.1398-9995.2000.00293.x

19. Shah A, Panjabi C. Allergic aspergillosis of the respiratory tract. European Respiratory Review. 2014; 23(131):8–29. doi: doi.org/10.1183/09059180.00007413

20. Agarwal R, Chakrabarti A, Shah A, et al. Allergic bronchopulmonary aspergillosis: review of literature and proposal of new diagnostic and classification criteria. Clinical & Experimental Allergy. 2013; 43(8):850–73. doi: 10.1111/cea.12141

21. Denning DW, Pashley C, Hartl D, et al. Fungal allergy in asthma–state of the art and research needs. Clinical and Translational Allergy. 2014; 4(1):14. doi: 10.1186/2045-7022-4-14

22. Jiang N, Xiang L. Allergic bronchopulmonary aspergillosis misdiagnosed as recurrent pneumonia. Asia Pacific Allergy. 2020; 10(3). doi: 10.5415/apallergy.2020.10.e27

23. Zhang M, Gao J. Clinical Analysis of 77 Patients with Allergic Bronchopulmonary Aspergillosis in Peking Union Medical College Hospital. Zhongguo Yi Xue Ke Xue Yuan Xue Bao. 2017; 39(3). doi: 10.3881/j.issn.1000-503X.2017.03.009

24. Cook PC, Brown SL, Houlder EL, et al. Mgl2+ cDC2s coordinate fungal allergic airway type 2, but not type 17, inflammation in mice. Nature Communications. 2025; 16(1). doi: 10.1038/s41467-024-55663-3

25. Liégeois M, Bai Q, Fievez Let al. Airway Macrophages Encompass Transcriptionally and Functionally Distinct Subsets Altered by Smoking. American Thoracic Society. 2022; 67(2). doi: 10.1165/rcmb.2021-0563OC

26. Tippalagama R, Singhania A, Dubelko P, et al. HLA-DR Marks Recently Divided Antigen-Specific Effector CD4 T Cells in Active Tuberculosis Patients. The Journal of Immunology. 2021; 207(2):523–33. doi: 10.4049/jimmunol.2100011

27. Cibrián D, Sánchez-Madrid F. CD69: from activation marker to metabolic gatekeeper. European Journal of Immunology. 2017; doi: 47(6):946–53.10.1002/eji.201646837

28. Kiazyk S, Fowke K. Loss of CD127 expression links immune activation and CD4+ T cell loss in HIV infection. Trends in Microbiology. 2008; 16(12). doi: 10.1016/j.tim.2008.08.011

29. Dunham RM, Cervasi B, Brenchley JM, et al. CD127 and CD25 expression defines CD4+ T cell subsets that are differentially depleted during HIV infection. The Journal of Immunology. 2008; 180(8). doi: 10.4049/jimmunol.180.8.5582

30. Bryant N, Muehling LM. T-cell responses in asthma exacerbations. Annals of Allergy, Asthma & Immunology. 2022; 129(6):709–18. doi: 10.1016/j.anai.2022.07.027

31. Hammad H, Lambrecht BN. The basic immunology of asthma. Cell. 2021; 184(6):1469–85. doi: 10.1016/j.cell.2021.02.016

32. Zarobkiewicz MK, Wawryk-Gawda E, Kowalska W, et al. γδ T Lymphocytes in Asthma: a Complicated Picture. Archivum Immunologiae et Therapiae Experimentalis. 2021;69(69). doi: 10.1007/s00005-021-00608-7

33. Weng N-P, Akbar AN, Goronzy J. CD28™ T cells: their role in the age-associated decline of immune function. Trends in Immunology. 2009; 30(7):306–12. doi: 10.1016/j.it.2009.03.013

34. Mou D, Espinosa J, Lo DJ, et al. CD28 Negative T Cells: Is Their Loss Our Gain? American Journal of Transplantation. 2014; 14(11):2460–6. doi: 10.1111/ajt.12937

35. Dua B, Tang W, Watson R, et al. Myeloid dendritic cells type 2 after allergen inhalation in asthmatic subjects. Clinical & Experimental Allergy. 2014; 44(7):921–9. doi: 10.1111/cea.12297

36. Dua B, Watson R, Guavreau G, et al. Myeloid and plasmacytoid dendritic cells in induced sputum after allergen inhalation in subjects with asthma. Journal of Allergy and Clinical Immunology. 2010; 126(1). doi: 10.1016/j.jaci.2010.04.006

37. Cella M, Engering A, Pinet V, et al. Inflammatory stimuli induce accumulation of MHC class II complexes on dendritic cells. Nature. 1997;388(6644):782–7. doi: 10.1038/42030

38. Reis E Sousa C. Dendritic cells in a mature age. Nature Reviews Immunology. 2006; 6(6):476–83. doi: 10.1038/nri1845

39. Neefjes J, Jongsma MLM, Paul P, et al. Towards a systems understanding of MHC class I and MHC class II antigen presentation. Nature Reviews Immunology. 2011; 11(12):823–36. doi: 10.1038/nri3084

40. Colombo SAP, Brown SL, Hepworth MR, et al. Comparative phenotype of circulating versus tissue immune cells in human lung and blood compartments during health and disease. Discovery Immunology. 2023; 2(1). doi: 10.1093/discim/kyad009

41. Vroman H, Hendriks RW, Kool M. Dendritic Cell Subsets in Asthma: Impaired Tolerance or Exaggerated Inflammation? Frontiers in Immunology. 2017; 8. doi: 10.3389/fimmu.2017.00941

42. Yun Y, Kanda A, Kobayashi Y, et al. Increased CD69 expression on activated eosinophils in eosinophilic chronic rhinosinusitis correlates with clinical findings. Allergology International. 2020; 69(2). doi: 10.1016/j.alit.2019.11.002

43. Hartnell A, Robinson D, Kay A, et al. CD69 is expressed by human eosinophils activated in vivo in asthma and in vitro by cytokines. Immunology. 1993; 80(2). doi: PMC1422202

44. Lucey DR, Nicholson-Weller A, Weller PF. Mature human eosinophils have the capacity to express HLA-DR. Proceedings of the National Academy of Sciences. 1989; 86(4):1348–51. doi: 10.1073/pnas.86.4.1348

45. Marc-Malovrh M, Camlek L, Škrgat S, et al. Elevated eosinophils, IL5 and IL8 in induced sputum in asthma patients with accelerated FEV1 decline. Respiratory Medicine. 2020; 162. doi: 10.1016/j.rmed.2020.105875

46. GINA. Pocket Guide For Asthma Management and Prevention. https://ginasthma.org/wp-content/uploads/2023/07/GINA-2023-Pocket-Guide-WMS.pdf Date last updated: July 2023. Date last accessed: May 12 2025

47. Huang K, Li F, Wang X, et al. Clinical and cytokine patterns of uncontrolled asthma with and without comorbid chronic rhinosinusitis: a cross-sectional study. Respiratory Research. 2022;23(1). doi: 10.1186/s12931-022-02028-3

48. Marriott H, Duchesne M, Moitra S, et al. Upper Airway Alarmin Cytokine Expression in Asthma of Different Severities. Journal of Clinical Medicine. 2024; 13(13):3721. doi: 10.3390/jcm13133721

49. Ma D, Muñoz X, Ojanguren I, et al. Increased TGFβ1, VEGF and IFN-γ in the Sputum of Severe Asthma Patients With Bronchiectasis. Archivos De Bronconeumologia. 2024; 60(11). doi: 10.1016/j.arbres.2024.05.036

50. Schwantes EA, Manthei DM, Denlinger LC, et al. Interferon gene expression in sputum cells correlates with the Asthma Index Score during virus-induced exacerbations. Clinical & Experimental Allergy. 2014; 44(6):813–21. doi: 10.1111/cea.12269

51. Hansel TT, Tunstall T, Trujillo-Torralbo M-B, et al. A Comprehensive Evaluation of Nasal and Bronchial Cytokines and Chemokines Following Experimental Rhinovirus Infection in Allergic Asthma: Increased Interferons (IFN-γ and IFN-λ) and Type 2 Inflammation (IL-5 and IL-13). EBioMedicine. 2017; 19:128–38. doi: 10.1016/j.ebiom.2017.03.033

52. Ray A, Kolls JK. Neutrophilic Inflammation in Asthma and Association with Disease Severity. Trends in Immunology. 2017; 38(12):942–54. doi: 10.1016/j.it.2017.07.003

53. Li Y, Wang W, Lv Z, et al. Elevated Expression of IL-33 and TSLP in the Airways of Human Asthmatics In Vivo: A Potential Biomarker of Severe Refractory Disease. The Journal of Immunology. 2018; 200(7):2253–62. doi: 10.4049/jimmunol.1701455

54. Calderon AA, Dimond C, Choy DF, et al. Targeting interleukin-33 and thymic stromal lymphopoietin pathways for novel pulmonary therapeutics in asthma and COPD. European Respiratory Review. 2023; 32(167):220144. 10.1183/16000617.0144-2022

55. Mcgrenaghan E, Smith AF. Airway management research: what problem are we trying to solve? Anaesthesia. 2019; 74(6):704–7. doi: 10.1111/anae.14563

56. Schwarz C, Eschenhagen P, Schmidt H, et al. Antigen specificity and cross-reactivity drive functionally diverse anti–Aspergillus fumigatus T cell responses in cystic fibrosis. Journal of Clinical Investigation. 2023; 133(5). Doi: 10.1172/JCI161593

57. Moon H-G, Eccles JD, Kim S-J, et al. Complement C1q essential for aeroallergen sensitization via CSF1R+ conventional dendritic cells type 2. Journal of Allergy and Clinical Immunology. 2023; 152(5):1141–52.e2. doi: 10.1016/j.jaci.2023.07.016

58. Massanari M, Holgate ST, Busse WW, et al. Effect of omalizumab on peripheral blood eosinophilia in allergic asthma. Respiratory Medicine. 2010; 104(2):188–96. doi: 10.1016/j.rmed.2009.09.011

59. Prazma CM, Bel EH, Price RG, et al. Oral corticosteroid dose changes and impact on peripheral blood eosinophil counts in patients with severe eosinophilic asthma: a post hoc analysis. Respiratory Research. 2019; 20(1). doi: 10.1186/s12931-019-1056-4

60. Franco P, Pereira G, Giavina-Bianchi P, et al. Inhaled corticosteroid use and its implication in peripheral eosinophil level. Journal of Allergy and Clinical Immunology. 2020; 145(2):AB28.

61. Humbert M, Taillé C, Mala L, Le Gros V, Just J, Molimard M. Omalizumab effectiveness in patients with severe allergic asthma according to blood eosinophil count: the STELLAIR study. European Respiratory Journal. 2018; 51(5):1702523. Doi: 10.1183/13993003.02523-2017

62. Fu Y, Wang J, Zhou B, et al. An IL-9–pulmonary macrophage axis defines the allergic lung inflammatory environment. Science Immunology. 2022; 7(68). doi: 10.1126/sciimmunol.abi9768

63. Yamada H, Yamaguchi M, Yamamoto K, et al. Eotaxin in induced sputum of asthmatics: relationship with eosinophils and eosinophil cationic protein in sputum. Allergy. 2000; 55(4):392–7. doi: 10.1034/j.1398-9995.2000.00474.x

64. Xu J, Jiang F, Nayeri F, et al. Apoptotic eosinophils in sputum from asthmatic patients correlate negatively with levels of IL-5 and eotaxin. Respiratory Medicine. 2007; 101(7):1447–54. doi: 10.1016/j.rmed.2007.01.026

65. Erpenbeck VJ, Hohlfeld JM, Volkmann B, et al. Segmental allergen challenge in patients with atopic asthma leads to increased IL-9 expression in bronchoalveolar lavage fluid lymphocytes. Journal of Allergy and Clinical Immunology. 2003; 111(6):1319–27. Doi: 10.1067/mai.2003.1485

66. Shimbara A, Christodoulopoulos P, Soussi-Gounni A, et al. IL-9 and its receptor in allergic and nonallergic lung disease: Increased expression in asthma. Journal of Allergy and Clinical Immunology. 2000; 105(1):108–15. doi: 10.1016/s0091-6749(00)90185-4

67. Pizzichini E, Pizzichini M, Efthimiadis A, et al. Measurement of inflammatory indices in induced sputum: effects of selection of sputum to minimize salivary contamination. European Respiratory Journal. 1996; 9(6):1174–80. doi: 10.1183/09031936.96.09061174

