## Supplementary figures and images for "Activation status of immune cells in the airway is a defining feature of severe fungal asthma"

### Supplementary Figure 1

**Supplementary Figure 1**

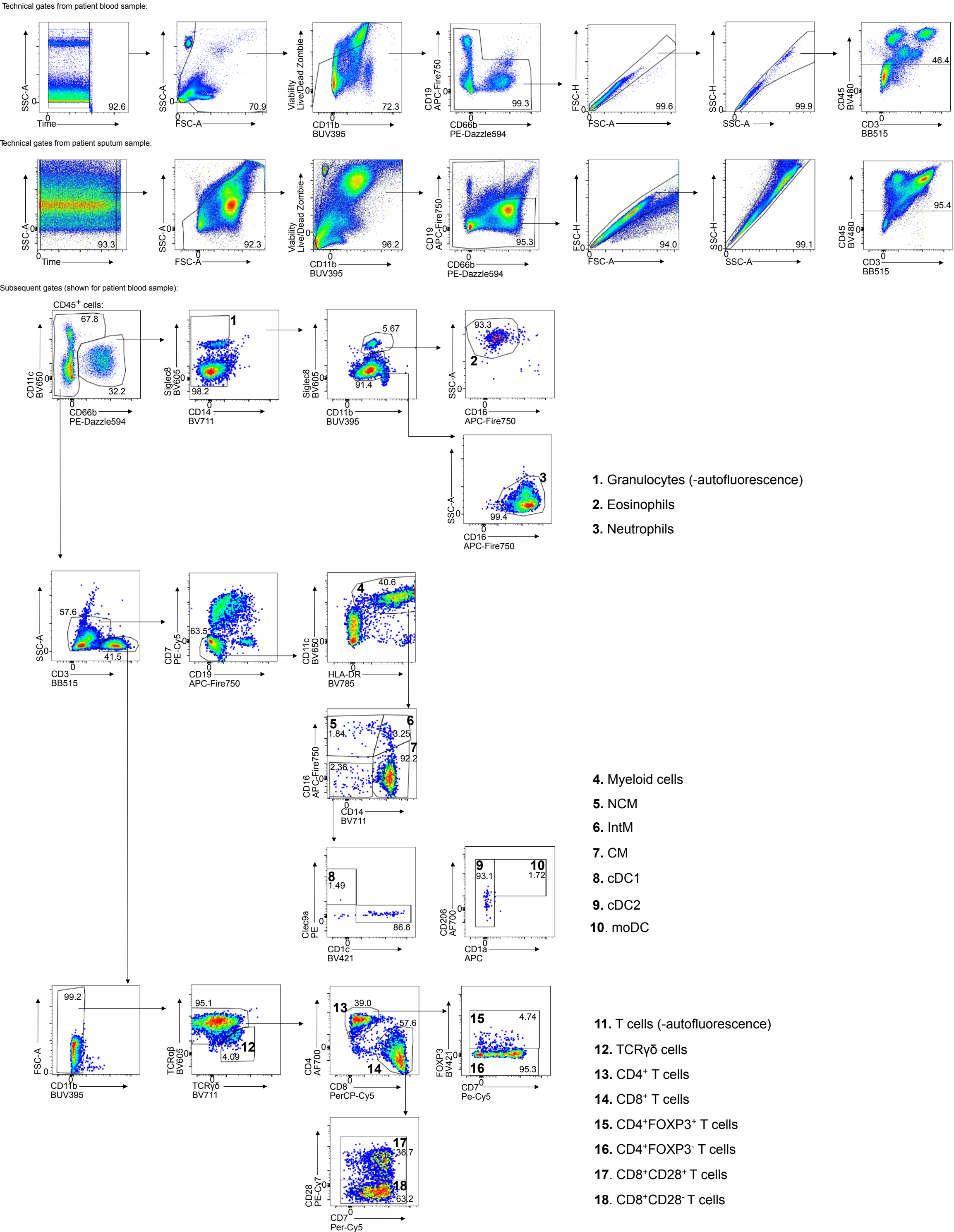

### Supplementary Figure 2

**Supplementary Figure 2**

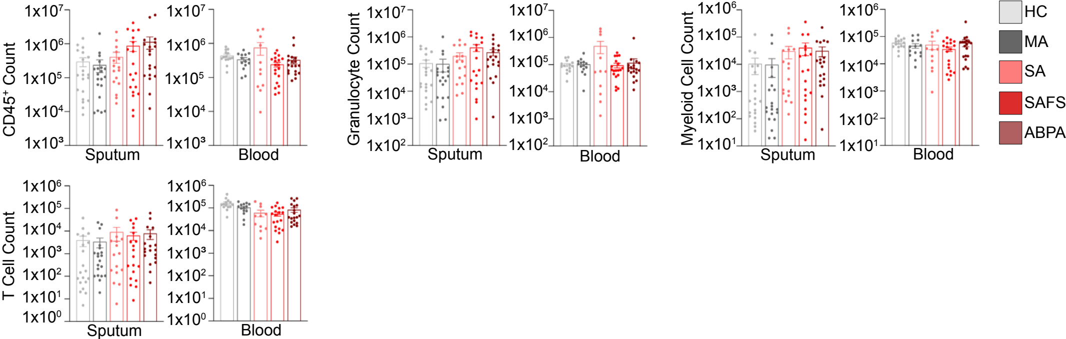
