## Supplementary Table 1 for "Activation status of immune cells in the airway is a defining feature of severe fungal asthma"

| Marker | Fluorochrome | Clone | Supplier |
|---|---|---|---|
| CD45 | BV480 | HI30 | BD Biosciences |
| CD3 | BB515 | UCHT1 | BD Biosciences |
| TCRγδ | BV711* | 11F2 | BD Biosciences |
| Siglec-8 | BV605* | 837535 | BD Biosciences |
| CD11b | BUV395 | ICRF44 | BD Biosciences |
| CD7 | PE-Cy5 | CD7-6B7 | BioLegend |
| CD66b | PE-Dazzle594 | G10F5 | BioLegend |
| CD11c | BV650 | Bu15 | BioLegend |
| HLA-DR | BV785 | L243 | BioLegend |
| TCRαβ | BV605* | IP26 | BioLegend |
| FOXP3 | BV421* | 206D | BioLegend |
| CD4 | AF700* | RPA-T4 | BioLegend |
| CD8 | PerCP-Cy5 | RPA-T8 | BioLegend |
| CD19 | APC-Fire750* | HIB19 | BioLegend |
| CD16 | APC-Fire750* | 3G8 | BioLegend |
| CD14 | BV711* | M5E2 | BioLegend |
| CD206 | AF700* | 15-2 | BioLegend |
| CLEC9a | PE* | 8F9 | BioLegend |
| CD1a | PE-Cy7* | HI149 | BioLegend |
| CD69 | APC | FN50 | BioLegend |
| CD127 | PE* | A019D5 | BioLegend |
| CD28 | PE-Cy7* | 37.51 | BioLegend |
| CD1c | BV421* | L161 | BD Biosciences |
| Live/Dead | Zombie | | ThermoFisher |

* = Fluorochrome shared by two markers for tandem gating strategy.
