## Supplementary Table 2 for "Activation status of immune cells in the airway is a defining feature of severe fungal asthma"

| Population | Marker | Association |
| --- | --- | --- |
| Eosinophils | CD69 | Activation |
| | HLA-DR | Maturation |
| CD4+FOXP3- T cells | CD69 | Activation, tissue residency |
| | HLA-DR | Activation |
| | CD127 | Conventional T cells |
| CD8+ T cells | CD69 | Activation, tissue residency |
| | HLA-DR | Activation |
| | CD28 | Exhaustion |
| | CD127 | Memory |
| cDC2s | HLA-DR | Activation |
| | CD206 | Maturation |
