## Supplementary Table 3 for "Activation status of immune cells in the airway is a defining feature of severe fungal asthma"

| Tissue | Population | Parameter | Exclusion criteria applied? | Exclusion criteria | Number of samples (HC) | Number of samples (MA) | Number of samples (SA) | Number of samples (SAFS) | Number of samples (ABPA) |
|---|---|---|---|---|---|---|---|---|---|
| **Sputum** | Eosinophils | %CD45 | N | n/a | 20 | 20 | 16 | 19 | 19 |
| | | Count | N | | | | | | |
| | | gMFI Siglec8 | Y | <400 cells | 6 | 6 | 9 | 11 | 12 |
| | | gMFI HLA-DR | Y | | | | | | |
| | | gMFI CD69 | Y | | | | | | |
| | Neutrophils | %CD45 | N | n/a | 20 | 20 | 16 | 19 | 19 |
| | CD4+ T cells | %CD45 | N | n/a | 20 | 20 | 16 | 19 | 19 |
| | CD4+ FOXP3+ T cells | %CD45 | N | n/a | 20 | 20 | 16 | 19 | 19 |
| | | % CD4+ | Y | <400 cells | 7 | 5 | 8 | 10 | 10 |
| | CD4+ FOXP3- T cells | %CD45 | N | n/a | 20 | 20 | 16 | 19 | 19 |
| | | % CD4+ | Y | <400 cells | 7 | 5 | 8 | 10 | 10 |
| | | HLA-DR MFI | Y | | | | | | |
| | | CD127 MFI | Y | | | | | | |
| | | CD69 MFI | Y | | | | | | |
| | CD8+ T cells | %CD45 | N | n/a | 20 | 20 | 16 | 19 | 19 |
| | | gMFI CD28 | Y | <300 cells | 7 | 7 | 8 | 9 | 12 |
| | | gMFI HLA-DR | Y | | | | | | |
| | | gMFI CD127 | Y | | | | | | |
| | | gMFI CD69 | Y | | | | | | |
| | CD8+ CD28+ T cells | %CD45 | N | n/a | 20 | 20 | 16 | 19 | 19 |
| | | % CD8 | Y | <300 cells | 7 | 7 | 8 | 9 | 12 |
| | CD8+ CD28- T cells | %CD45 | N | n/a | 20 | 20 | 16 | 19 | 19 |
| | | % CD8 | Y | <300 cells | 7 | 7 | 8 | 9 | 12 |
| | TCRγδ T cells | %CD45 | N | n/a | 20 | 20 | 16 | 19 | 19 |
| | Classical Monocytes | %CD45 | N | n/a | 20 | 20 | 16 | 19 | 19 |
| | Non-classical Monocytes | %CD45 | N | n/a | 20 | 20 | 16 | 19 | 19 |
| | Intermediate monocytes | %CD45 | N | n/a | 20 | 20 | 16 | 19 | 19 |
| | cDC1s | %CD45 | N | n/a | 20 | 20 | 16 | 19 | 19 |
| | cDC2s | %CD45 | N | n/a | 20 | 20 | 16 | 19 | 19 |
| | | gMFI HLA-DR | Y | <340 cells | 8 | 6 | 9 | 11 | 17 |

| | | | | | | | | | |
|---|---|---|---|---|---|---|---|---|---|
| | | gMFI CD206 | Y | | | | | | |
| | moDCs | %CD45 | N | n/a | 20 | 20 | 16 | 19 | 19 |
| **Blood** | Eosinophils | %CD45 | N | n/a | 14 | 14 | 11 | 19 | 19 |
| | | gMFI Siglec8 | Y | <210 cells | 14 | 13 | 10 | 19 | 18 |
| | | gMFI HLA-DR | Y | | | | | | |
| | | gMFI CD69 | Y | | | | | | |
| | Neutrophils | %CD45 | N | n/a | 14 | 14 | 11 | 19 | 19 |
| | CD4+ T cells | %CD45 | N | n/a | 14 | 14 | 11 | 19 | 19 |
| | CD4+ FOXP3+ T cells | %CD45 | N | n/a | 14 | 14 | 11 | 19 | 19 |
| | | % CD4+ | N | | | | | | |
| | CD4+ FOXP3- T cells | %CD45 | N | n/a | 14 | 14 | 11 | 19 | 19 |
| | | % CD4+ | N | | | | | | |
| | | HLA-DR MFI | N | | | | | | |
| | | CD127 MFI | N | | | | | | |
| | | CD69 MFI | N | | | | | | |
| | CD8+ T cells | %CD45 | N | n/a | 14 | 14 | 11 | 19 | 19 |
| | | gMFI CD28 | N | | | | | | |
| | | gMFI HLA-DR | N | | | | | | |
| | | gMFI CD127 | N | | | | | | |
| | | gMFI CD69 | N | | | | | | |
| | CD8+ CD28+ T cells | %CD45 | N | n/a | 14 | 14 | 11 | 19 | 19 |
| | | % CD8 | N | | | | | | |
| | CD8+ CD28- T cells | %CD45 | N | n/a | 14 | 14 | 11 | 19 | 19 |
| | | % CD8 | N | | | | | | |
| | TCRγδ T cells | %CD45 | N | n/a | 14 | 14 | 11 | 19 | 19 |
| | Classical Monocytes | %CD45 | N | n/a | 14 | 14 | 11 | 19 | 19 |
| | Non-classical Monocytes | %CD45 | N | n/a | 14 | 14 | 11 | 19 | 19 |
| | Intermediate monocytes | %CD45 | N | n/a | 14 | 14 | 11 | 19 | 19 |
| | cDC1s | %CD45 | N | n/a | 14 | 14 | 11 | 19 | 19 |
| | cDC2s | %CD45 | N | n/a | 14 | 14 | 11 | 19 | 19 |
| | | gMFI HLA-DR | Y | <100 cells | 14 | 14 | 10 | 15 | 16 |
| | | gMFI CD206 | Y | | | | | | |
| | moDCs | %CD45 | N | n/a | 14 | 14 | 11 | 19 | 19 |
